# Surveillance by age-class and prefecture for emerging infectious febrile diseases with respiratory symptoms, including COVID-19

**DOI:** 10.1101/2020.04.11.20061697

**Authors:** Tomoaki Ueno, Junko Kurita, Tamie Sugawara, Yoshiyuki Sugishita, Yasushi Ohkusa, Hirokazu Kawanohara, Miwako Kamei

## Abstract

**Object:** The COVID-19 outbreak emerged in late 2019 in China, expanding rapidly thereafter. Even in Japan, epidemiological linkage of transmission was probably lost already by February 18, 2020. From that time, it has been necessary to detect clusters using syndromic surveillance.

**Method:** We identified common symptoms of COVID-19 as fever and respiratory symptoms. Therefore, we constructed a model to predict the number of patients with antipyretic analgesics (AP) and multi-ingredient cold medications (MIC) controlling well-known pediatric infectious diseases including influenza or RS virus infection. To do so, we used the National Official Sentinel Surveillance for Infectious Diseases (NOSSID), even though NOSSID data are weekly data with 10 day delays, on average. The probability of a cluster with unknown febrile disease with respiratory symptoms is a product of the probabilities of aberrations in AP and MIC, which is defined as one minus the probability of the number of patients prescribed a certain type of drug in PS compared to the number predicted using a model. This analysis was conducted prospectively in 2020 using data from October 1, 2010 through 2019 by prefecture and by age-class.

**Results:** The probability of unknown febrile disease with respiratory symptom cluster was estimated as less than 60% in 2020.

**Discussion:** The most severe limitation of the present study is that the proposed model cannot be validated. A large outbreak of an unknown febrile disease with respiratory symptoms must be experienced, at which time, practitioners will have to “wing it”. We expect that no actual cluster of unknown febrile disease with respiratory symptoms will occur, but if it should occur, we hope to detect it.

## Introduction

The novel *Coronavirus* COVID-19 outbreak emerged in late 2019, subsequently expanding to 93,090 laboratory-confirmed cases and 3,018 deaths reported by WHO as of March 6 [1]. In Japan, excluding cruise ships, 349 cases including asymptomatic cases were confirmed, from which six patients died. However, most have been diagnosed as mild cases as of March 6, 2020 [2]. The epidemiological linkage of transmission was probably untraceable already in Japan as of February 18, 2020, as it was in Wuhan. At that time, cluster identification was necessary, but rapid testing was unavailable. Therefore, physicians have been unable to diagnose potential patients especially if they have mild symptoms. Therefore, syndromic surveillance is expected to be as useful as traditional surveillance based on diagnosis by physicians. The present study examines Prescription Surveillance (PS) for detection of some clusters of emerging infectious febrile disease with respiratory symptom including unknown febrile disease with respiratory symptoms [3, 4] as syndromic surveillance.

In actuality, PS has been operating in Japan since 2009. The system provides nationwide syndromic surveillance by the Japan Medical Association, Japan Pharmaceutical Association, and EM Systems Co. Ltd. It has been reporting the estimated number of influenza and chicken pox patients and the numbers of patients prescribed antibiotic drugs, antipyretic analgesics (AP), and multi-ingredient cold medications (MIC) and antidiarrheal and intestinal drugs, based on prescriptions filled at pharmacies. The system estimates the number of patients every day, as inferred from the number of prescriptions by prefecture by age group. As of the end of January 2020, approximately ten thousand pharmacies were participating, collectively accounting for about 20% of all pharmacies nationwide. The estimated numbers of patients are presented on the dedicated web site the following morning [5]. It is the most reliable real-time precision and timely surveillance for influenza [6].

In Japan, National Official Sentinel Surveillance for Infectious Diseases (NOSSID) has been operating based on the related law [7]. For many common pediatric infectious diseases, 3,000 sentinel pediatric care facilities, which account for about one-tenth of all pediatric care facilities throughout Japan, report the number of cases each week by NOSSID. For influenza, 2,000 internal medicine care facilities have been added. The sentinel reports are regarded as reliable information because they are based on physicians’ diagnoses. Nevertheless, these reports are published weekly after a ten-day delay following diagnosis. For that reason, NOSSID probably cannot detect infectious diseases early. Moreover, because it is based on diagnoses, it cannot detect emerging or re-emerging diseases that cannot be diagnosed at medical institutions, at least in the initial phase of the outbreak.

The earlier study for detection of emerging infectious diseases using PS proposed a model that predicts the number of patients controlling calendar information and activity of common infectious diseases in NOSSID [8]. When it found unexplained increases in some type of drug, it reports an aberration. The model in this study is a modified version that addresses AP and MIC, corresponding to typical symptoms of COVID-19 patients [9]. The numbers of patients prescribed other types of drugs were used as additional control subjects.

## Methods

PS estimates the numbers of patients by multiplying the reciprocal of the pharmacy PS participation rate and the reciprocal of the proportion of prescriptions at external pharmacies in the prefecture by the total number of prescriptions issued in the prefecture. The numbers of patients prescribed anti-influenza virus drugs, anti-herpes virus drugs, antibiotic drugs, antipyretic analgesics (AP), multi-ingredient cold medications (MIC), and antidiarrheal and intestinal drugs have been recorded. Antibiotics are classifiable into five types: penicillin, cephem, macrolide, new quinolone, and others [5].

To predict the numbers of patients taking those drugs by well-known infectious diseases, we used data for influenza, RS virus infection, pharyngoconjunctival fever, group A streptococcal pharyngitis, gastrointestinal infections, varicella, hand, foot and mouth disease (HFMD), erythema infectiosum, exanthem subitum, pertussis, herpangina, mumps, and mycoplasma pneumonia from NOSSID.

First, from known infectious diseases and calendar information, we predicted the numbers of patients prescribed AP and MIC. The dependent variable was the number of patients prescribed drug *i* on day *t*. Explanatory variables were the NOSSID reported number of patients of disease *j*: the latest available data for day *t*. Because NOSSID publishes data of the prior week on Friday at noon, the latest available data are those of two weeks prior on Monday–Friday, and one week prior on Saturday and Sunday. Furthermore, explanatory variables included dummy variables of the epidemiological week, day of the week, holiday, and the day following a holiday, the day following two consecutive holidays, summer vacation (13–15 August), and the new year vacation (1–3 January). Moreover, the number of patients prescribed with drugs other than AP and MIC were included as explanatory variables.

Next, we calculated the probabilities of the numbers of patients prescribed a certain type of drug in PS comparison to the number predicted by the model. The probability of aberration for the drug was defined as one minus the probabilities of the numbers of patients prescribed a certain type of drug in PS compared with the number predicted by the model. The probability of a cluster with emerging diseases is the product of the probability of aberration in drugs corresponding to symptoms of the emerging diseases. For example, for COVID-19, because we presume that its typical symptoms are high fever and respiratory symptoms, we can presume that the probability of clusters with unknown febrile disease with respiratory symptoms is the product of the probabilities of aberrations in AP and MIC. This analysis, conducted prospectively in 2020, uses data from October 1, 2010 through 2019 by prefecture and by age class. Age classes were defined as 14 and younger, 15–65, and 65 years and older. We adopted 97.5% as the tentative criterion to detect clusters, which corresponds to a significance level of 5%.

## Results

Detected clusters of unknown febrile disease with respiratory symptoms based on the tentative criterion and its probability of aberration are shown in the table by age class. There were 17 clusters: 7 of children, 7 of adults, and 3 of elderly people. Clusters with probabilities of unknown febrile disease with respiratory symptoms that were higher than .99 were two: on February 5 among children in Mie prefecture and on January 7 among adults in the same prefecture.

**Table.** Detected clusters with unknown febrile disease with respiratory symptoms based on the tentative criterion and its probability of aberration

|  | Date |  | Prefecture | Probability |
| --- | --- | --- | --- | --- |
| Children | February | 5 | Mie | .9985045 |
|  | February | 7 | Nara | .9766988 |
|  | February | 8 | Mie | .9868521 |
|  | February | 10 | Aichi | .9895230 |
|  | February | 10 | Fukuoka | .9791710 |
|  | February | 18 | Nara | .9849889 |
|  | February | 25 | Aichi | .9846296 |
| Adults | January | 5 | Hiroshima | .9858211 |
|  | January | 6 | Iwate | .9757143 |
|  | January | 6 | Wakayama | .9766867 |
|  | January | 7 | Mie | .9901777 |
|  | January | 16 | Kumamoto | .9769740 |
|  | February | 3 | Chiba | .9785242 |
|  | February | 22 | Yamanashi | .9800284 |
| Elderly | January | 21 | Shizuoka | .9841036 |
|  | February | 18 | Fukushima | .9836578 |
|  | February | 21 | Osaka | .9852280 |

Note: This table presents detected aberrations of clusters with unknown febrile disease with respiratory symptoms based on a tentative criterion: .975.

The figure shows the number of patients with AP or MIC among children (upper panel) and adults (lower panel). The arrow indicates the date on which the probabilities of cluster were higher than 99%. It shows that AP or MIC in children or adults were not significantly higher than on other days.

**Figure.**
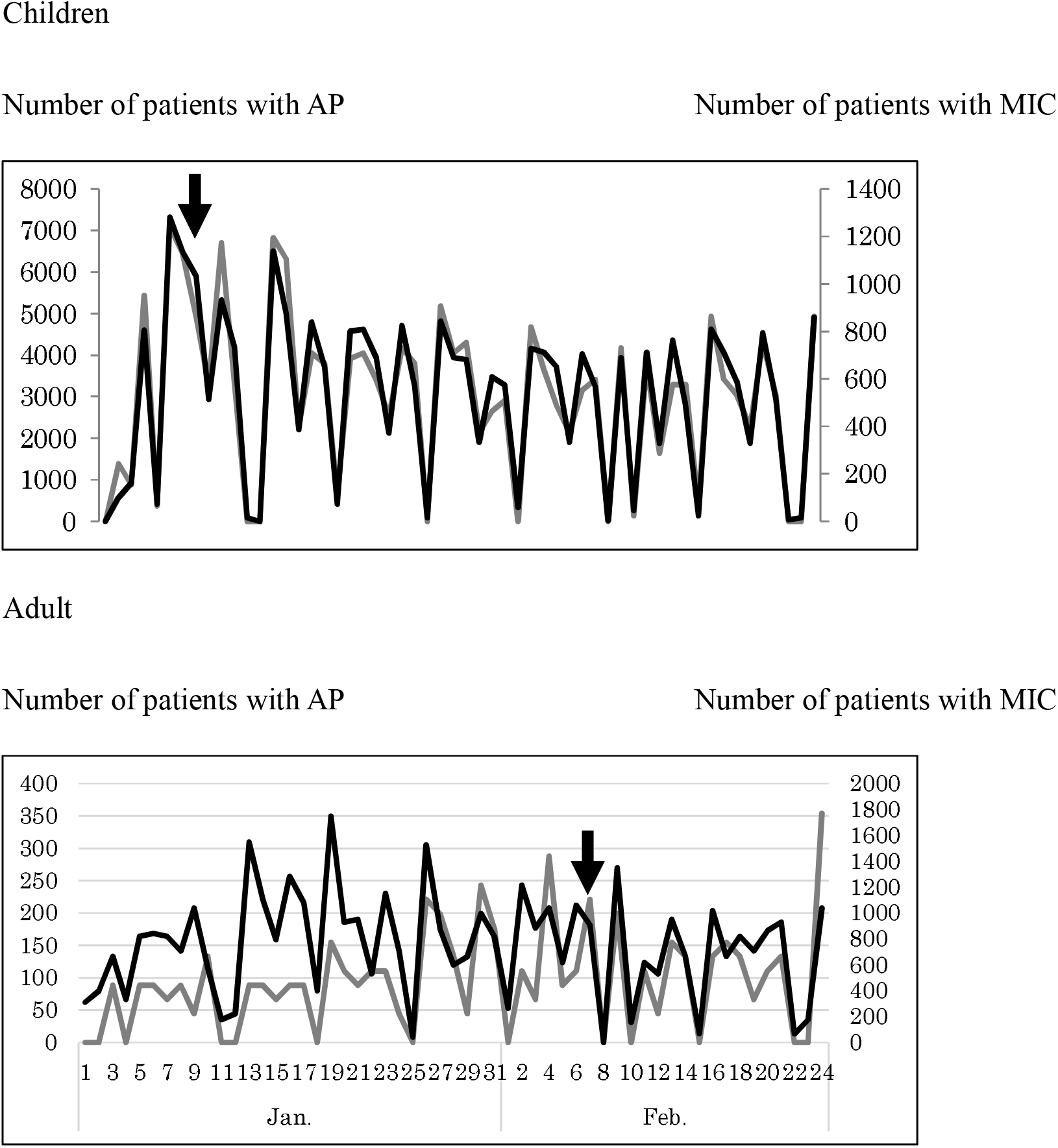
Number of patients with antipyretic analgesics (AP) and multi-ingredient cold (MIC) medications in 2020 among children and adults in Mie prefecture. Note: The upper panel presents the number of patients with AP orMIC medication for children. The lower panel shows the two drug types among adults in Mie prefecture. The scale for AP was measured at the left-hand side. The scale for MIC was measured at the right-hand side. Arrows show the detection date on which the probability of the cluster was greater than .99.

## Discussion

Results show 17 clusters of unknown febrile disease with respiratory symptoms for which the probabilities were higher than 97.5%. Only two clusters were detected for which probabilities were higher than 99%. Therefore, 97.5% was accepted as the criterion for moderate aberration; 99% was used as the criterion for high aberration. In addition, the lower aberration might be defined as higher than 90%.

However, it is noteworthy that the detected clusters were significantly more numerous in patients with unknown febrile disease with respiratory symptom. They might not be infected COVID-19. However, they were not with influenza, RS virus or other common pediatric infectious diseases.

We can use more real-time daily surveillance of infectious diseases other than PS and NOSSID, which were used for this study. For example, Nursery School Absenteeism Surveillance System ((N)SASSy) has been monitoring the health situation, diagnosed diseases or symptoms, of more than five million students or nursery school kids every day. It is a computerized system comprising a network of information shared among concerned organizations and individuals [10–12]. We can therefore use (N)SASSy as a form of syndromic surveillance in Japan. In short, all information related to children’s health conditions is integrated in (nursery) schools. It was developed by a research group headed by Dr. Ohkusa, one author of this paper. It has been funded by the MHLW since 2007. It has retained its copyright to the present day. Currently, it is operated by the Japanese Society of School Health.

Online Receipt Computer Advantage (ORCA) surveillance data compiled since 2010 have been provided for public use [13]. The data include estimated daily numbers of patients by prefecture or municipality in Japan for influenza, hand-foot-and-mouth disease (HFMD), erythema infectiosum (EI), pharyngoconjunctival fever (PCF), respiratory syncytial (RS) virus infection, measles, rubella, and insolation from ORCA, an electronic medical claim system (http://infect.orca.med.or.jp/) with data produced and then provided freely to medical facilities by the Japan Medical Association. As of the end of September in 2019, ORCA was used at 17,174 medical facilities. ORCA collected information from about 4218 medical facilities as of the end of September, 2019. As inferred from its 100 thousand medical facilities in total, its participation rate was about 4%. Actually, its precision was confirmed for influenza, which was very similar in PS at the nationwide. However precision for other diseases might be insufficient. Even so, because it has other information that is not included in PS, integration of ORCA surveillance and/or (N)SASSy might contribute to improvement of detection of COVID-19 or other emerging infectious diseases.

The most severe limitation of the present study is that the validity of the proposed model has not been verified. Verifying it requires that we experience a large outbreak of COVID-19. With such a system, practitioners would have to wing it during its first trial. Hopefully we expect to be able to detect any cluster of COVID-19 correctly if such a cluster were to exist. The prior day’s data of detected clusters have been published on the internet (http://prescription.orca.med.or.jp/syndromic/kanjyasuikei/index.php) since the beginning of March. To date, fortunately, we have no detection from this survey method.

The present study supports the author’s opinions, which are unrelated to any stance or policy of professionally affiliated bodies.

## Data Availability

Japan Ministry of Health, Labour and Welfare. Press Releases. (in Japanese)

https://www.mhlw.go.jp/stf/houdou/houdou_list_202003.html

## Acknowledgments

We acknowledge all pharmacies participating in PS.

